# SARS CoV-2 escape variants exhibit differential infectivity and neutralization sensitivity to convalescent or post-vaccination sera

**DOI:** 10.1101/2021.02.22.21252002

**Authors:** Alona Kuzmina, Yara Khalaila, Olga Voloshin, Ayelet Keren-Naus, Liora Bohehm, Yael Raviv, Yonat Shemer-Avni, Elli Rosenberg, Ran Taube

## Abstract

Towards eradicating COVID19, developing vaccines that induce high levels of neutralizing antibodies is a main goal. As counter measurements, viral escape mutants rapidly emerge and potentially compromise vaccine efficiency. Herein we monitored ability of convalescent or Pfizer-BTN162b2 post-vaccination sera to neutralize wide-type SARS-CoV2 or its UK-B.1.1.7 and SA-B.1.351 variants. Relative to convalescent sera, post-vaccination sera exhibited higher levels of neutralizing antibodies against wild-type or mutated viruses. However, while SARS-CoV2 wild-type and UK-N501Y were similarly neutralized by tested sera, the SA-N501Y/K417N/E484K variant moderately escaped neutralization. Significant contribution to infectivity and sensitivity to neutralization was attributed to each of the variants and their single or combined mutations, highlighting alternative mechanisms by which prevalent variants with either N501Y or E484K/K417N mutations spread. Our study validates the clinical significance of currently administered vaccines, but emphasizes that their efficacy may be compromised by circulated variants, urging the development of new ones with broader neutralization functions.

## INTRODUCTION

One year into the outbreak of the Severe Acute Respiratory Syndrome Coronavirus 2 (SARS-CoV-2) in Wuhan China, it is clear that the pandemic is here to stay for the foreseeable future, likely resulting in substantial addition of morbidity and mortality (Zhou et al., 2020; Zhu et al., 2020). Currently COVID-19 has spread worldwide, infecting 109M people and resulting in 2,41M deaths. The viral Spike (S) protein consists of an N-terminal (S1) subunit that mediates receptor binding, and a C-terminal (S2) subunit responsible for virus-cell membrane fusion (Wrapp et al., 2020). Upon viral entry into cells, the receptor-binding domain (RBD) of spike engages the primary receptor, human angiotensin converting enzyme 2 (hACE2) (Letko et al., 2020), followed by cleavage at a Furin cleavage site by host cell proteases, TMPRSS2, TMPRSS4 or endosomal cathepsins, facilitating S2-dependent fusion of viral and host-cell membranes (Hoffmann et al., 2020; Zang et al., 2020).

Neutralizing antibodies (nAb) that target SARS-CoV-2-ACE2 engagement, arise mainly against the RBD (Alsoussi et al., 2020; Brouwer et al., 2020; Gaebler et al., 2021; Group et al., 2020; Rogers et al., 2020; Wu et al., 2020). As such, characterizing the epitopes that are recognized by these nAb and defining the natural viral mutants within the spike, are important to outline SARS-CoV-2 escape variants, and should predict the proper use of antibody-based countermeasures. In late 2020, several COVID-19 vaccines have been introduced and are currently being administrated globally. The leading two are mRNA-based vaccines developed by Pfizer (BNT162b2) and Moderna (mRNA-1273) (Anderson et al., 2020; Krammer, 2020; Polack et al., 2020; Walsh et al., 2020). Both vaccines target the viral spike and potentially elicit high neutralizing titers that efficiently block infection and prevent disease progression (Anderson et al., 2020; Baden et al., 2021). Recent work shows that the Pfizer vaccine elicits similar neutralization patterns as convalescent sera, providing disease protection (Polack et al., 2020; Walsh et al., 2020). However, as the virus persists and exhibits relatively moderate to high mutation rate, it evades the immune response by developing escape variants with resistance to sera from recovered patients or vaccinated individuals. Two significant variants, which display multiple mutations in their spike have emerged and spread worldwide at a high pace. Variant B.1.1.7 was first detected in the United Kingdom (UK) and appears to possess an increased transmissibility (Kemp et al., 2021a; Kemp et al., 2020; Kemp et al., 2021b; Korber et al., 2020). Along with the dominant D614G mutation, which improves viral fitness and transmission (Hou et al., 2020; Korber et al., 2020; Plante et al., 2020), additional mutations in its spike protein were acquired, thereby improving viral transmissibility and escape from nAb. Among them, a 69/70 N-terminal deletion is associated with reduced sensitivity to neutralization, as well as the N501Y, A570D, and P681H mutations. Importantly, the N501Y and P681H are located within the RBD and furin-cleavage site, respectively. N501Y interacts in a tighter manner with ACE2, enabling the virus to expand its host range and increase its infectivity (Chan et al., 2020; Wrapp et al., 2020; Xie et al., 2020). B.1.351 is a second variant that was reported in South Africa (SA). It shares the Y501N mutation, and also includes nine changes in the S protein (Giandhari et al., 2020). One cluster in N-terminal domain includes four substitutions and a deletion (L18F; D80A; D215G; delta 242-244 and R246I), while a second cluster contains three substitutions in the RBD (K417N; E484K and N501Y). Early reports have indicated that while the RBD mutation N501Y in the B.1.1.7 strain does not compromise post vaccine serum neutralization (Shi et al., 2021), E484K partly impairs neutralization (Greaney et al., 2021), potentially compromising effectiveness of neutralizing sera and currently available vaccines (Baum et al., 2020; Ku et al., 2021; Liu et al., 2021; Muik et al., 2021; Wang et al., 2021a; Wang et al., 2021c; Weisblum et al., 2020; Wibmer et al., 2021a; Wu et al., 2021a),(Andreano et al., 2020; Xie et al., 2021). In addition, to these two major variants, additional mutations of the same nature have been also described in the new - Brazil-P1 variant. In such a rapidly evolving pandemic, it is highly important to determine the nature of neutralization sensitivity of emerging escape variants to sera from convalescent or vaccinated individuals. Here we assessed the neutralizing activity of 25 sera samples from convalescent or vaccinated individuals against the wild type SARS-CoV-2 and its B.1.1.7 or B.1.351 variants. We also determined the contribution of the RBD mutations that appear in each of the two variants and their combinations, to sera neutralization and viral infectivity. Our results show that sera from the second-dose vaccinated individuals exhibited an eleven-fold increase in neutralization activity relative to convalescent sera. Moreover, RBD mutations, N501Y, K417N and E484K (or their combinations) escaped neutralization activity of tested sera at varying extents. Effects of these mutations were also documented to the end-point viral infectivity. While N501Y containing variants did not escape neutralization, their infectivity rates were high relative to wild-type SARS-CoV-2. In contrast, the E484K and to a lesser extent K417N containing variants, conferred escape from neutralizing antibodies (nAb). However, they all exhibit infectivity values similar to wild-type SARS-CoV-2, exhibiting only a 2-fold increase in their infectivity. Importantly, the SA-N501Y/K417N/E484K variant exhibited both high infectivity and successfully escapes neutralization. Finally, while escape from sera neutralization was detected in both convalescent and post-vaccination sera, the substantial increase in neutralization activity due to vaccination boost, highlighted the clinical significance of vaccination.

## RESULTS

### Wild type SARS-CoV-2 is neutralized to a different extent by convalescent or post-vaccination sera

We initially tested the ability of a panel of convalescent sera drawn from recovered COVID-19 patients, which exhibited moderate to severe disease (n=10), or sera drawn from Pfizer BNT162b2 post-vaccinated individuals (3 weeks post dose #1; n=5, and 9-11 days post dose #2; n=10) to neutralize wild-type SARS-CoV-2 spike (S) pseudotyped lentivirus following infection of HEK-ACE2 cells. Our results show that all sera samples exhibited neutralizing activity against the wild-type SARS-CoV-2 (**Figure 2**). Significantly, sera from vaccinated participants who received the second dose, exhibited an average two-fold increased levels of neutralizing titers, with mean IC_50_ dilution of 99,000, relative to sera drawn 3 weeks post the first dose of vaccination – mean IC_50_ dilution of 51,300. Importantly, we documented eleven-fold increase in neutralizing activity of the post-vaccination sera relative to sera drawn from convalescent sera - mean IC_50_ dilution of 8700. A 6-fold increase in neutralization activity was obtained when sera from the first vaccination dose (3 weeks post-vaccination) was compared to convalescent sera. These data imply that the second dose of vaccination is critical to obtain optimized neutralization titers against wild type SARS-CoV-2 (**Figure 2)**. Preferably, COVID-19 recovered patients should be also vaccinated to boost their nAb titers. Moreover, our neutralization results correlate well with measurements of levels of specific SARS-CoV-2 IgG, presented in Table 1 and 2.

**Table 1:** Convalescent human sera.

| serum | Days post symptoms<br>onset | ELISA IgG<br>Liasonx | IC-50 dilution |
| --- | --- | --- | --- |
| P1 | 63 | 8 | 100000 |
| P3 | 56 | 37 | 100000 |
| P4 | 21 | 0 | 79.36 |
| P5 | 49 | 6 | 250 |
| P6 | 28 | 21 | 833.3 |
| P7 | 21 | 20 | 10000 |
| P8 | 63 | 68 | 25000 |
| P9 | 56 | 23 | 20000 |
| P10 | 49 | 17 | 10000 |
| P11 | 49 | 11 | 833.3 |

**Table 2:** Post vaccination human sera.

| serum | Days post<br>dose #1 | Days post dose #2 | ELISA IgG<br>Liasonx | IC-50 dilution |
| --- | --- | --- | --- | --- |
| V.1.1 | 21 |  | 79 | 50000 |
| V.1.2 | 21 |  | 35 | 33333.3 |
| V.1.3 | 21 |  | 63 | 33333.3 |
| V.1.4 | 21 |  | 109 | 40000 |
| V.1.5 | 21 |  | 119 | 100000 |
| V.2.2 |  | 9 | 259 | 200000 |
| V.2.3 |  | 9 | 254 | 100000 |
| V.2.4 |  | 9 | 190 | 100000 |
| V.2.5 |  | 9 | 156 | 64000 |
| V.2.6 |  | 7 | 400 | 128000 |
| V.2.7 |  | 8 | 223 | 83333.3 |
| V.2.8 |  | 11 | 326 | 83333.3 |
| V.2.9 |  | 9 | 341 | 83333.3 |
| V.2.10 |  | 8 | 211 | 50000 |
| V.2.11 |  | 10 | 222 | 100000 |

**Figure 1:**
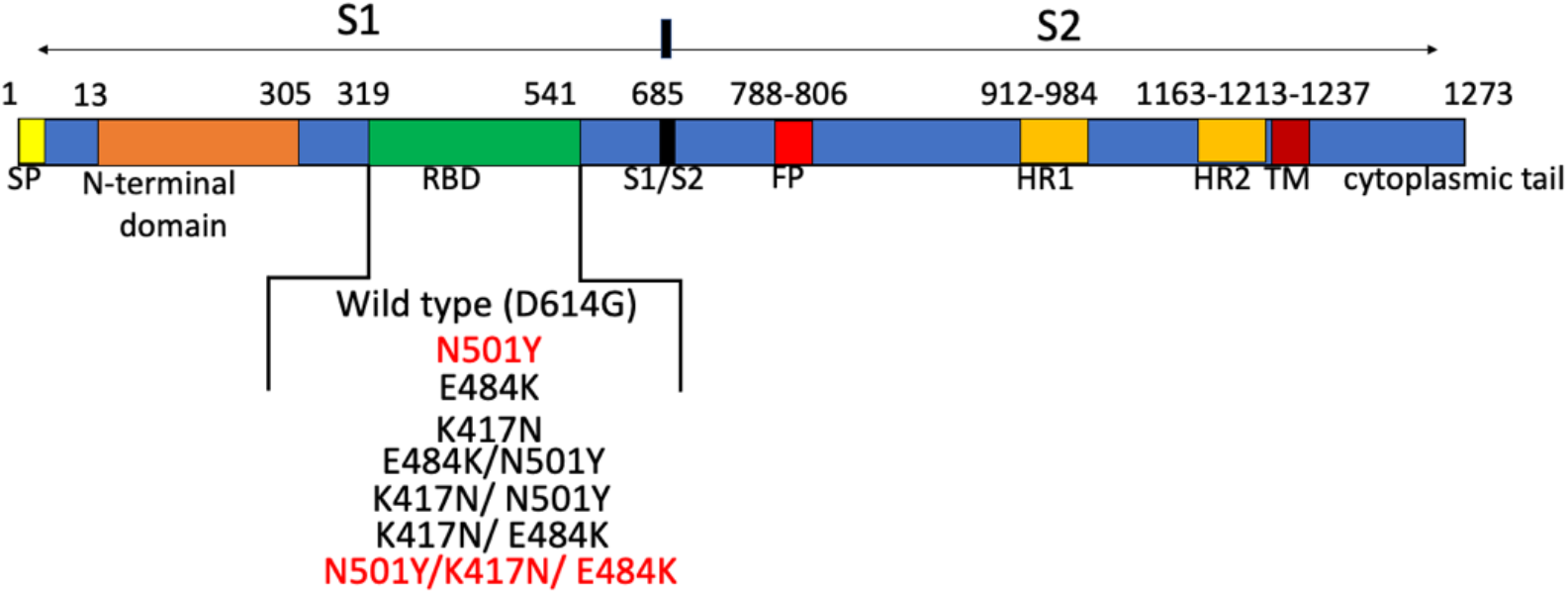
Schematic organization of the Spike (S) protein of SARS CoV-2. Schematic organization of the spike (S) protein with defined domains, and mutants that were introduced within the RBD spike gene.

**Figure 2:**
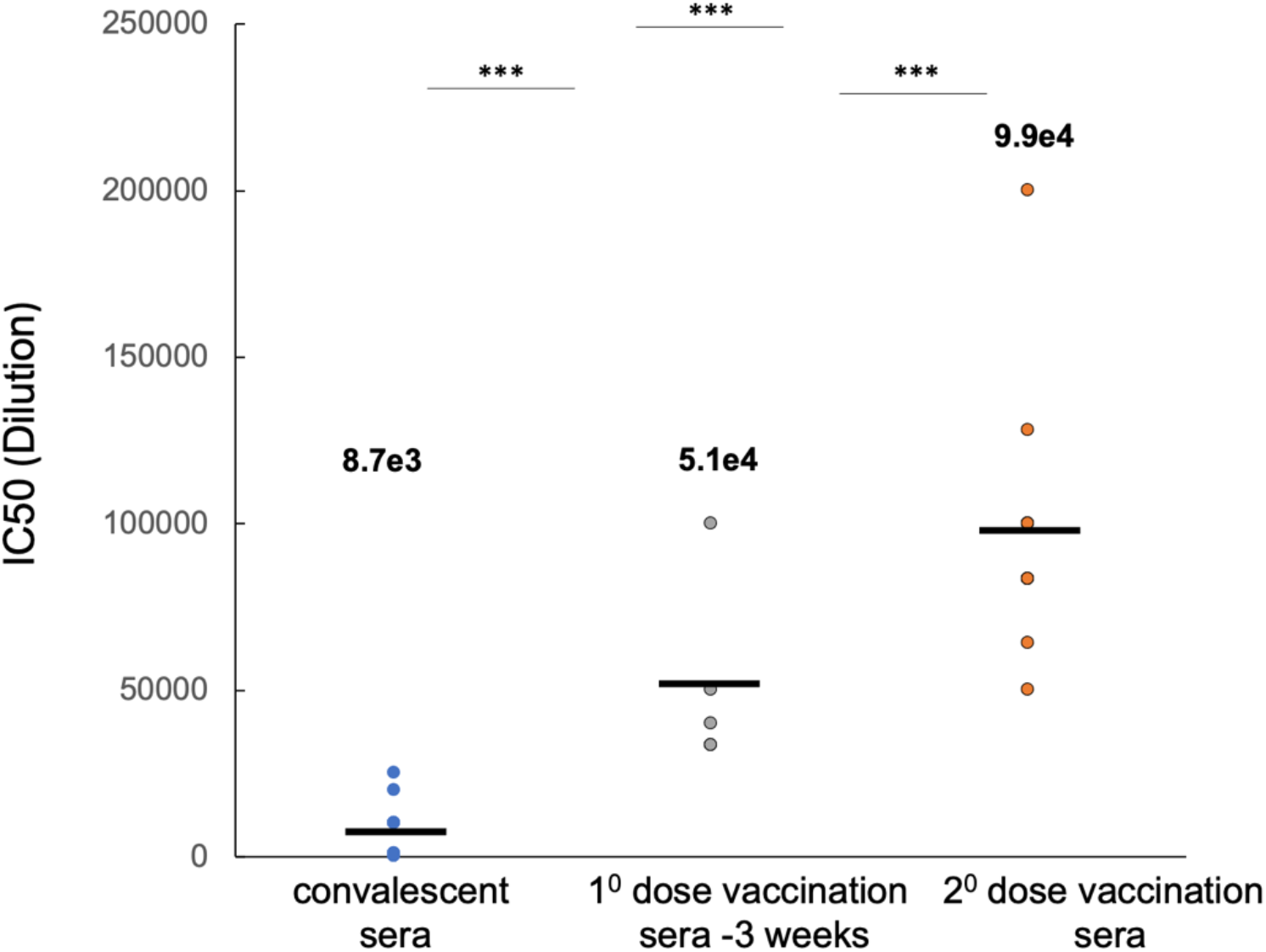
Human convalescent or post-vaccination sera neutralize wild type. SARS-CoV-2 Reporter lentiviruses pseudotyped with either wild type SARS-CoV-2 spike variants were used to infect HEK293T-ACE2 stable cells, in the presence of decreasing dilutions sera drawn from convalescent or post vaccinated individuals (3 weeks following dose #1, or 9-11 days post dose #2). 48hr post infection, cells were harvested and their luciferase readings were monitored according to the manufacturer protocol (Promega). Sera neutralizing activity was calculated at serial dilutions, relative to infected cells with no sera added. IC50 is defined as the dilution that mediated 50% neutralization relative to control infected cells with no sera. Results are mean of average of two independent experiments. Black bars represent geometric mean of IC50 values, indicated at the top. Statistical significance was determined using one tailed t-test ***<0.001.

### Neutralization of the UK-N501Y and SA-N501Y/K417N/E484K RBD mutants by convalescent or post-vaccination sera

The recently emerged SARS-CoV-2 RBD mutants emerging in the UK and SA variants exhibit higher transmissibility, thus rapidly spread worldwide. We thus monitored the ability of our sera samples to neutralize the N501Y RBD mutation found in the UK strain, as well as the SA-N501Y/K417N/E484K mutations of the SA strain. IC_50_ values were monitored relative to the wild-type strain (**Figure 3A+B**). Our data show that convalescent serum neutralized the wild-type strain with an average of IC_50_ dilution of 8700. Interestingly, the UK-N501Y strain was also neutralized by the convalescent sera, exhibiting only a 1.5-fold decrease in neutralization activity, relative to the wild-type strain. In contrast, the SA-N501Y/K417N/E484K variant exhibited higher resistance to neutralization by convalescent sera, with an average 6.8-fold decrease relative to the wild type strain (**Figure 3A**). Moreover, there was a rather wide distribution of IC_50_ ratio values between the wild-type and the SA, reaching up to 80-fold. Upon vaccination, levels of neutralizing antibodies increased, confirming our data in **Figure 2**. Second dose post vaccinated sera, exhibited similar neutralization capabilities, both to the wild type and the UK-N501Y strain, implying the high efficiency of the vaccine against the UK strain (**Figure 3B**). However, the ability of post-vaccinated sera to neutralize the SA-N501Y/K417N/E484K variant decreased 6.8-fold relative to wild type viral strain (**Figure 3B**). In this case, the distribution of IC_50_ ratios between samples was relatively small. We conclude that the SA-N501Y/K417N/E484K escape variant exhibits an increased resistance to neutralization against both convalescent and Pfizer vaccinated sera.

**Figure 3:**
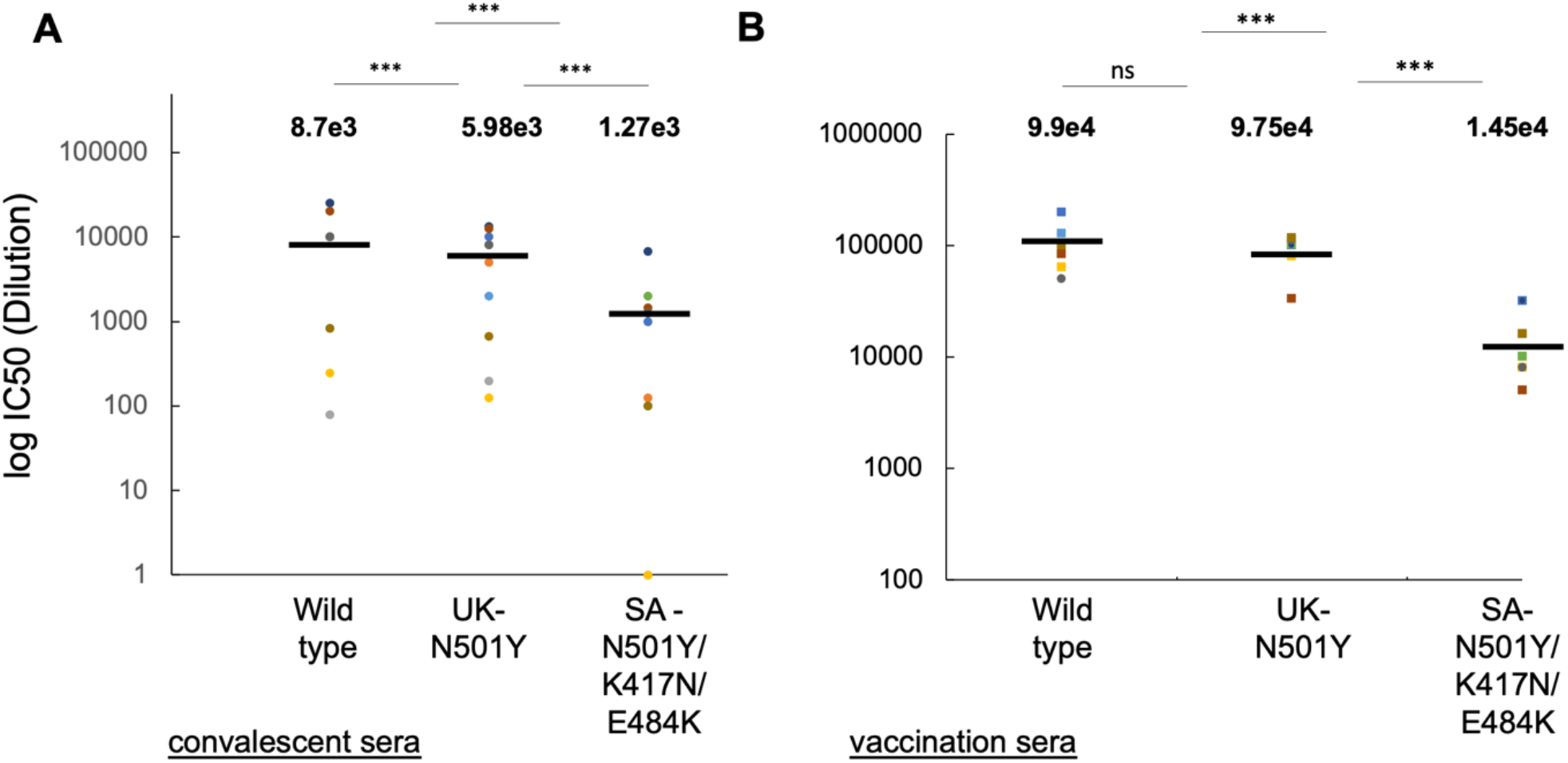
Neutralization of the UK-N501Y and SA-N501Y/K417N/E484K variants by convalescent and post-vaccination sera. Reporter single round reporter lentiviruses pseudotyped with either wild type or mutant SARS-CoV-2 spike UK-N501Y or SA-N501Y/K417N/E484K variants were used to infect HEK293T-ACE2 stable cells, in the presence of decreasing dilutions of either convalescent serum (**A**) n=10); or sera drawn from vaccinated individuals’ dose 2 (**B**) (n=10). 48hr post infection, cells were harvested and their luciferase readings were monitored according to the manufacturer protocol (Promega). Serum neutralizing activity was calculated at serial dilutions, relative to infected cells with no sera added. IC50 is defined as the dilution that mediated 50% neutralization relative to control infected cells with no sera. Results are mean of average of two independent experiments. Horizontal black bars represent geometric mean IC50 values, indicated at the top. Statistical significance was determined using one tailed t-test ***P<0.001.

### RBD mutants of UK and SA SARS-CoV2 variants affect viral infectivity

We next tested the relative infectivity of our RBD mutants using spike pseudotyped lentivirus. HEK-293T-ACE2 cells were infected with equal MOI of the UK-N501Y and SA-N501Y/K417N/E484K variants, as well as with pseudotyped lentiviruses with different single or combined mutations of the relevant residues - K417N, E484K, N501Y/E484K, N501Y/K417N and K417N/E484K (**Figure 4A**). Our results show that relative to the wild-type SARS-CoV-2, set to 1, the infectivity of the UK-N501Y strain significantly increased up to 9-fold. The infectivity of the SA-N501Y/K417N/E484K strain further exhibited a small increase, reaching to 12-fold relative to the wild type strain. We also evaluated the contribution of the other RBD mutations. Interestingly, the E484K single mutant virus exhibited only a 2-fold increase in its infectivity relative to the wild type strain. Similarly, the K417N single mutant virus and the K417N**/**E484K double mutant virus also showed only a two-fold increase in their infectivity relative to the wild type strain. Interestingly, viral variants where the N501Y mutation was attached to either K417N or E484K (N501K/K417N or N501K/E484K) exhibited high infectivity rates, closer to those exhibited by the UK-N501Y variant. These results highlight the contribution of the contribution of N501Y mutation to viral infectivity (**Figure 4A**), suggesting an alternative mechanism of spread for the UK variants.

**Figure 4:**
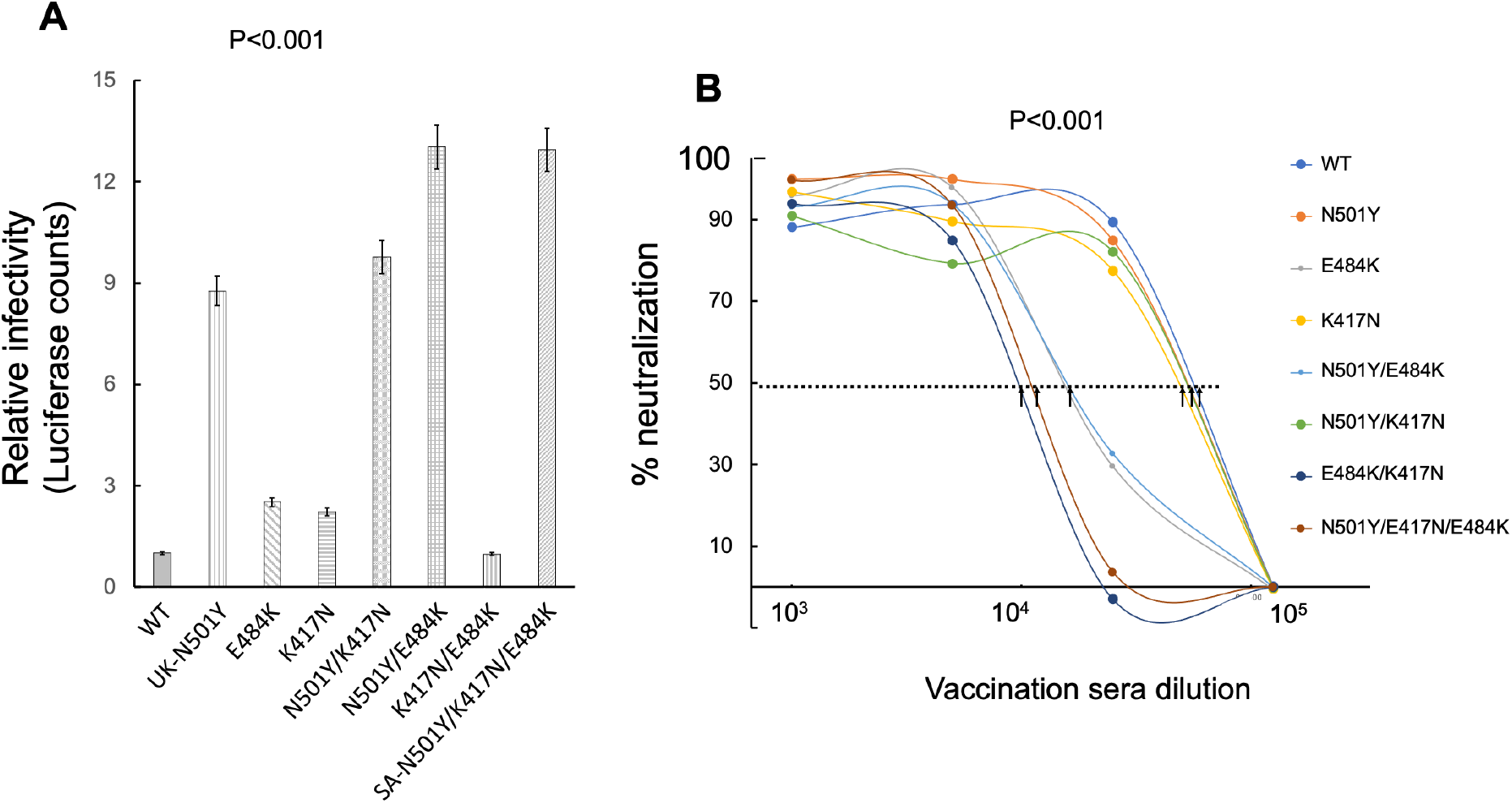
Sensitivity of SARS-CoV-2 variants and their combined spike mutants to neutralization by post-vaccination sera. A. N501Y RBD mutation is key for viral infection of SARS-CoV-2 variants Reporter single round lentiviruses pseudotyped with either wild type or the indicated mutant SARS-CoV-2 spike variants were used to infect HEK293T-ACE2 stable cells at equal MOI. 48hr post infection, cells were harvested and their luciferase readings were monitored according to the manufacturer protocol (Promega). Infectivity is corrected to MOI values based on p24 counts measured by ELISA. Bar graphs show mean values ± SD error bars of three independent experiments. Measured statistical significance was calculated between experiments by a two-tailed Student’s t test - P≤ 0.001. B. E484K and K417N RBD mutations are key for neutralizing SARS-CoV-2 variants Reporter lentiviruses pseudotyped with either wild type or the indicated mutant SARS-CoV-2 spike UK-N501Y, or SA-N501Y/K417N/E484K variants were used to infect HEK293T-ACE2 stable cells, in the presence of decreasing dilutions of a mixed serum from samples drawn from vaccinated individuals dose 2 (n=10). 48hr post infection, cells were harvested and their luciferase readings were monitored according to the manufacturer protocol (Promega). Sera % neutralizing activity was calculated at serial dilutions, relative to infected cells with no sera added. IC50 is defined as the dilution that mediated 50% neutralization relative to control infected cells with no sera. Arrows indicate IC50 values obtained for each mutant. IC50 values for WT, K417N, N501Y, N501Y/K417N were around 1.5e5. IC50 for K417N, E484K and N501Y/E484K were around 6.5e5. And finally, IC50 values for N501Y/K417N/E484K, E484K/K417N were approximately 1e4. Results are mean of average of two independent experiments. Black arrows represent mean IC50 dilution values. Measured statistical significance was calculated between experiments by a two-tailed Student’s t test - P≤ 0.001.

### Neutralization sensitivity of RBD mutants to post-vaccination sera

We also tested the ability of vaccinated sera (a mixture of samples from the second dose) to neutralize our different viral variants, in a spike pseudotyped neutralization assay (**Figure 4B**). We confirm above results showing that the UK-N501Y variant was similarly neutralized by vaccinated sera as the wild type viral strain. K417N and N501Y/K417N viral mutants also were similarly neutralized as the wild type strain. However, viruses that displayed the E484K and N501Y/E484K mutations led to a moderate resistance to neutralization by vaccinated sera. Finally, the K417N/E484K and the SA-N501Y/K417N/E484K variants exhibited the highest resistant to vaccinated sera, emphasizing the role of E484K in nAb neutralizing escape (**Figure 4B**). These results highlight the contribution of the contribution of E484K and K417N mutations to escape from neutralization.

## DISCUSSION

The induction of neutralizing antibodies (nAb) that target the RBD of the SARS-CoV-2 spike (S) protein through vaccination is a main route towards complete eradication of the COVID19 pandemic. However, the virus evades the immune response selective pressure and escape-mutants that resist neutralization emerge and may compromise the efficacy of administered vaccines. Indeed, new SARS-CoV-2 variants, UK-B.1.1.7 and SA-B.1.351, recently appeared and are rapidly spread worldwide. In this study, we examined whether the currently administered Pfizer vaccine efficiently neutralizes SARS-CoV-2 escape variants. We also determined which of the mutations within the RBD, or their combinations, are responsible for the potential neutralization resistance and for enhanced infectivity. Our results show that the UK-N501Y variant is highly infectious, as it exhibited a nine-fold increase in viral infectivity, relative to the wild-type SARS-CoV-2 (**Figure 4A**). Moreover, when N501Y mutation was joint by other mutations - K417N (N501Y/K417N), E484K (N501Y/E484K), or N501Y/K417N/E484K - an enhanced ten to fourteen-fold infectivity rates were documented relative to the wild type virus. On the other end of the infectivity spectrum, viruses with K417N or E484K single mutations, or the combined K417N/E484K all exhibited infectivity rates that were similar to the wild type strain (two-fold increase). Unlike infectivity, escape from neutralization seemed to be dependent on the E484K, and to a lesser extent on the K417N mutations. Viruses displaying E484K alone or the combined N501Y/E484K successfully escaped neutralization. Furthermore, the E484K/K417N as the SA-N501Y/K417N/E484K viruses optimally escaped neutralization by post-vaccination sera relative to wild-type SARS-CoV-2 (**Figure 4B**). Based on these results, we suggest that there are alternative mechanisms for elevated transmissibility and spread of SARS-CoV-2 that come into play within its main escape variants. While N501Y is a key mutation that potentially enhances association of the virus with its ACE2 receptor, thus increases infectivity and spread, the E484K and K417N mutations drive viral spread primarily through neutralization escape. It is likely that viral strains that harbor combinations of all of these relevant mutations circulate and affect disease spread. These variants have the potential to further evolve and mutate, leading to a loss of neutralization activity as partly seen in the SA strain.

Our data also concludes that the administrated Pfizer vaccine is moderately compromised against SA-N501Y/K417N/E484K escape variant. Average decrease in neutralization function of the vaccinated sera samples against this variant were 6.8-fold relative to wild-type SARS CoV-2. This result is only partly aligned with recent conclusions from Pfizer, claiming that their vaccine is almost similarly efficient against the South African variant as the wild type SARS CoV-2 (Xie et al., 2021). Moreover, a report from Moderna also documented a 6.4-fold reduction in neutralizing the SA-B.1.351 variant by its vaccine. However, their conclusion was that such a reduction in neutralizing titers against are not clinically significant (Wu et al., 2021b). However, the clinical significance of a 6-fold reduced neutralization activity of convalescent or vaccinated sera against the SA strain remains to be determined. Other reports align better with our data, reporting that the SA variant, or its combined mutations, confer neutralization resistance from specific monoclonal antibodies and vaccinated sera (Wang et al., 2021b; Wang et al., 2021d; Wibmer et al., 2021b).

Nonetheless, we show that while in vaccination sera the distribution from average neutralization titers was relatively narrow, in convalescent sera there was a rather wide range of distribution from the average titers between samples, and some samples were unable to neutralize the SA virus at all (**Figure 3**). These results also imply that vaccination is clinically significant as it boosts the levels of neutralizing antibodies. We suggest that special attention needs to be taken to avoid the SA variant spread as it may moderately resist vaccination sera. Nevertheless, our results are conclusive regarding the need to obtain the two doses of the vaccine, as levels of neutralizing antibodies in post vaccination sera were higher compared to convalescent sera, urging the need to be vaccinated even after recovering from COVID19.

## Data Availability

All data are included within the manuscript

## ACKNOWLEDGMENTS

This work was supported by the Israeli Mistry of Science and Technology (MOST; grant #3-16897) and the Israel Science Foundation (ISF; Research Grant Application no. 755/17).

## AUTHOR CONTRIBUTIONS

A.K and R.T conceived the study and analyzed the data. A.K., Y.H. and O.V. performed experiments and analyzed the data. A.N., Y.S., L.B., Y.R., and E.R helped with obtaining sera samples. E. R., and R.T submitted the Helsinki request. R.T. wrote the manuscript.

## DECLERATION OF INTRESTS

The authors have no conflicts of interest to declare.

Sera samples drawn from ten (n=10) COVID19 recovered individuals were collected at the indicated time points post onset of disease symptoms and screened by SARS-CoV-2 IgG Liasonx ELISA for specific SARS-CoV-2 IgG. Similarly, sera samples drawn from individuals that were vaccinated by the BNT162b2 Pfizer COVID19 vaccine were also collected at 3 weeks post first dose (V.1; n=5), or 9 days post the second dose (V.2; n=10). The different sera were then monitored by neutralization assays against wild type or spike mutated SARS-CoV S pseudotyped lentivirus.

## METHODS

## Spike mutant generation

QuikChange Lightening Site-Directed Mutagenesis kit was used to generate amino acid substitutions in the pCDNA Spike plasmid, following the manufacturer’s instructions (Agilent Technologies, Inc., Santa Clara, CA).

## Sera sample collection, pseudotyped lentivirus production and neutralization assays

Sera was collected from COVID19 convalescent or from vaccinated individuals, 21 days post first dose, or 9-11 days post second dose. Collected sera are summarized below in Table 1 and 2. The study was conducted in compliance with the ethical principles of the Declaration of Helsinki and approved by the intuitional ethics committee. To confirm total SARS-CoV-2 IgG in the collected sera, specific IgG levels were determined by Liasonx ELISA. HEK-ACE2 stable cells were infected with luciferase reporter lentivirus, pseudotyped with either wild type spike of SARS CoV-2 or different combinations of its S protein mutants.

Pseudotyped viruses were generated in HEK293T cells. Briefly, LTR-PGK luciferase lentivector was transfected into cells together with other lentiviral expression plasmids coding for either Gag, Pol Tat Rev, and the corresponding wild type or mutate S envelopes. Transfections were done in a 10cm format, as previously described(Krasnopolsky et al., 2020).

Pseudotyped viruses were incubated with decreasing dilutions of the tested sera or without sera as a control, for 1 hr at 37°C. HEK-ACE2 cells were then infected in a 96 well format for 12 hr. 72hr post infection, cells were harvested and analyzed by luciferase assay according to the manufacturer protocol (Promega). Measurements were performed in duplicate and used to calculate IC50 – 50% inhibitory sera neutralizing concentration.

